# Investigating the likely association between genetic ancestry and COVID-19 manifestations

**DOI:** 10.1101/2020.04.05.20054627

**Authors:** Ranajit Das, Sudeep D. Ghate

**Author notes:** All correspondence to Dr. Ranajit Das, Yenepoya Research Centre, Yenepoya (Deemed to be University), University Road, Deralakatte, Mangalore 575018, Karnataka, India.

## Abstract

**Background:** The novel coronavirus: severe acute respiratory syndrome coronavirus 2 (SARS-CoV-2) has spread rapidly throughout the world leading to catastrophic consequences. However, SARS-CoV-2 infection has shown discernible variability across the globe. While in some countries people are recovering relatively quickly, in others, recovery times have been comparatively longer and number of individuals succumbing to it are high. This variability in coronavirus disease 2019 (COVID-19) susceptibility is suggestive of a likely association between the genetic-make up of affected individuals modulated by their ancestry and the severity of COVID-19 manifestations.

**Objective:** In this study, we aimed to evaluate the potential association between an individual’s genetic ancestry and the extent of COVID-19 disease presentation employing Europeans as the case study. In addition, using a genome wide association (GWAS) approach we sought to discern the putative single nucleotide polymorphism (SNP) markers and genes that may be likely associated with differential COVID-19 manifestations by comparative analyses of the European and East Asian genomes.

**Method:** To this end, we employed 10,215 ancient and modern genomes across the globe assessing 597,573 SNPs obtained from the databank of Dr. David Reich, Harvard Medical School, USA to evaluate the likely correlation between European ancestry and COVID-19 manifestations. Ancestry proportions were determined using qpAdm program implemented in AdmixTools v5.1. Pearson’s correlation coefficient (r) between various ancestry proportions of European genomes and COVID-19 death/recovery ratio was calculated and its significance was statistically evaluated. Genome wide association study (GWAS) was performed in PLINK v1.9 to investigate SNPs with significant allele frequency variations among European and East Asian genomes that likely correlated with differential COVID-19 infectivity.

**Results:** We found significant positive correlation (*r*=0.58, *P*=0.03) between West European hunter gatherers (WHG) ancestral fractions and COVID-19 death/recovery ratio for data as of 5^th^ April 2020. This association discernibly amplified (*r*=0.77, *P*=0.009) upon reanalyses based on data as of 30^th^ June 2020, removing countries with small sample sizes and adding those that are a bridge between Europe and Asia. Using GWAS we further identified 404 immune response related SNPs by comparing publicly available 753 genomes from various European countries against 838 genomes from various Eastern Asian countries. Prominently, we identified that SNPs associated with immune-system related pathways such as interferon stimulated antiviral response, adaptive and innate immune system and IL-6 dependent immune responses show significant differences in allele frequencies [Chi square values (≥1500; *P*≈0)] between Europeans and East Asians.

**Conclusion:** So far, to the best of our knowledge, this is the first study investigating the likely association between host genetic ancestry and COVID-19 severity. These findings improve our overall understanding of the putative genetic modifiers of COVID-19 clinical presentation. We note that the development of effective therapeutics will benefit immensely from more detailed analyses of individual genomic sequence data from COVID-19 patients of varied ancestries.

## Introduction

The novel coronavirus, severe acute respiratory syndrome coronavirus 2 (SARS-CoV-2), was officially identified on 7^th^ January 2020 in Wuhan, China [1]. Since then, it has spread rapidly throughout the world leading to devastating consequences. Notably, COVID-19, caused by SARS-CoV-2 has shown significant worldwide variability, especially in terms of the death/recovery ratio that pertains to a ratio of the number of deaths caused by it to numbers of infected individuals who have recovered within the same time interval. While affected individuals in some parts of the world have recovered relatively quickly with lower morbidities, those from various other regions appear to remain sick for longer with slower recovery times, more debilitating consequences and demonstrate relatively higher death/recovery ratios [2]. For instance, Verity *et al*. (2020) estimated an overall fatality ratio of 2·7% for COVID-19 cases reported outside of mainland China. For people with travel history to mainland China this ratio was 1.1% and those without any travel history to China the ratio was 3.6% [3]. This is suggestive of a likely population level genetic variation in terms of susceptibility to SARS-CoV-2 and COVID-19 manifestations.

A case in point in this context appears to be *ACE2* that encodes for angiotensin I converting enzyme 2 demonstrated to be the host receptor for the SARS-CoV-2 [4,5]. Recently, Cao *et al*. analysed 1700 variants in *ACE2* from The China Metabolic Analytics Project (ChinaMAP) and 1000 Genomes Project databases [6]. Their study revealed significant variation in allele frequencies among the populations assessed, notably among people of East Asian and European ancestries. They speculated that the differences in allele frequencies of *ACE2* coding variants, likely associated with its elevated expression may influence *ACE2* function and putatively impact the susceptibility, symptoms, and outcome of SARS-CoV-2 infection in evaluated populations. Additionally, a majority of the 15 expression quantitative trait loci (eQTL) variants they identified had discernibly higher allele frequencies among East Asian populations compared to Europeans, which may suggest differential susceptibility towards SARS-CoV-2 in these two populations under similar conditions. Stawiski *et al*. who performed a comprehensive analysis on >290,000 samples representing > 400 populations, also identified natural *ACE2* variants that can potentially alter the virus-host interaction and thus alter host susceptibility towards COVID-19 manifestations [7]. While some of these variants can potentially increase susceptibility for COVID-19, others were speculated to be protective and surmised to display decreased binding to SARS-CoV-2 spike protein and thus decrease susceptibility to infection.

In addition to *ACE2*, COVID-19 manifestations may also be modulated by several immune response modulating genes. Li *et al*. reviewed the interaction between coronaviruses and the innate immune systems of the hosts [8]. It was speculated that coronavirus infection can result in secretion of higher amounts of chemokines and cytokines such as interleukins (IL-1, IL-6, IL-8, IL-21), Tumor necrosis factor (TNF)-β and monocyte chemoattractant protein-1 (MCP-1)) in infected cells. Further, the hosts’ major antiviral machinery, largely composed of interferons (IFNs) can also act to restrain SARS-CoV-2 infection. Overall it may be speculated that variations in host genes involved it its antiviral response may be responsible for differential clinical presentation of COVID-19. Recently, genetic variability across major histocompatibility complex (MHC) class I genes [9] and apolipoprotein E *(APoE)* [10] have also been associated with severity of COVID-19 manifestations.

Further recent studies have shown that variants associated with immune responsiveness display significant population-specific signals of natural selection [11]. In particular, the authors showed that admixture with Neanderthals may have differentially shaped the immune systems in European populations, introducing novel regulatory variants, which may affect preferential responses to viral infections. European genomes were further modulated and distinguished owing to multiple waves of migration throughout their ancient past. We note here that while Neolithic European populations predominantly descended from Anatolian migrants, European ancestry shows a distinct West–East cline in indigenous West European hunter gatherers (WHGs) [12]. At the same time Eastern Europe was in the rudimentary stages of the formation of Bronze Age steppe ancestry, which later spread into Central and Northern Europe through East-West expansion [13]. Further, high genetic substructure between North-Western and South-Eastern hunter-gatherers has also been documented recently [14].

Ancestry specific variation in immune responsiveness and its potential genetic and evolutionary determinants are poorly understood especially in the context of SARS-CoV-2 infection and COVID-19 disease manifestations. In addition, despite its broad implications in identifying novel variants putatively associated with variation in host immune responses across different ancestries, only a handful of genome wide association study (GWAS) have been performed pertaining to viral infections, especially in the context of SARS-CoV-2 infection [15].

Here we aimed to unravel the potential association between host genetic ancestry and COVID-19 disease employing Europeans as the case study. In addition, using a GWAS approach we sought to discern the putative single nucleotide polymorphism (SNP)s, genes and pathways that may be potentially associated with differential COVID-19 infection by comparative analyses of the European and East Asian genomes. To the best of our knowledge, so far this is the first study to assess the relationship between host genetics and SARS-CoV-2 severity comparing individuals from distinct ancestries.

## Method

### Dataset

Genome data for the current study was obtained from the personal database of Dr. David Reich’s Lab Harvard Medical School, USA [16]. The final dataset comprised of 10,215 ancient and modern genomes across the globe assessing 597,573 single nucleotide polymorphisms (SNPs). File conversions and manipulations were performed using EIGENSTRAT (EIG) v7.2 [17] and PLINK v1.9 [18].

COVID-19 data was obtained from 2019 Novel Coronavirus COVID-19 Data Repository by The Center for Systems Science and Engineering, Johns Hopkins (JHU-CSSE) [2]. The data available in this web portal as of Sunday, 5^th^ April 2020, 6 PM was used in the primary analyses. To evaluate the robustness of our results, we repeated our analyses with the data available in Johns Hopkins CSSE web portal as of 30^th^ June 2020, 4:33 AM. Indian state-wise data was obtained from John Hopkins CSSE portal and confirmed through Government of India coronavirus portal as of 30^th^ June 2020 [19].

### Ancestry proportions in European genomes and COVID-19 manifestations

We employed whole genome data from ten European countries in the initial analyses (Table 1). During reanalysis, countries such as Norway, Czech Republic and Ukraine were removed due to smaller sample size (N<15). Instead, Eastern European countries such as Romania, Bulgaria, and those that bridge between Europe and Asia such as Georgia, Turkey and Armenia were included. Further, UK that did not provide officially updated COVID-19 recovery information was excluded during reanalysis. Furthermore, a larger sample size was employed for Italians (TSI) obtained from 1000 genomes project database (Table 2).

**Table 1:**
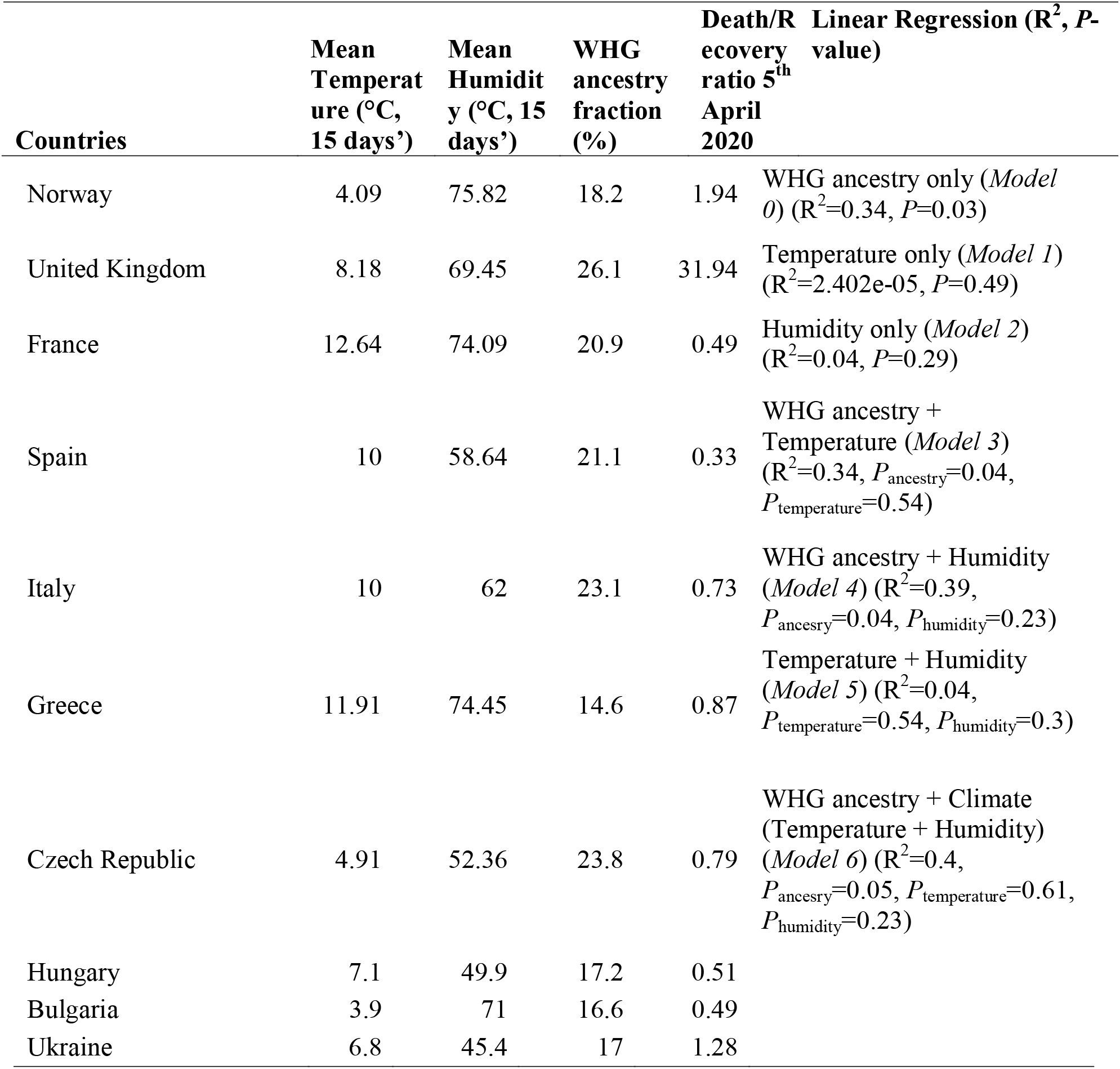
West European hunter gatherers (WHGs) ancestry fractions, mean temperature (15 days), mean humidity (15 days), and COVID-19 death/recovery ratios of 10 European countries employed in the study for initial analysis (data obtained for 5^th^ April 2020)

**Table 2:**
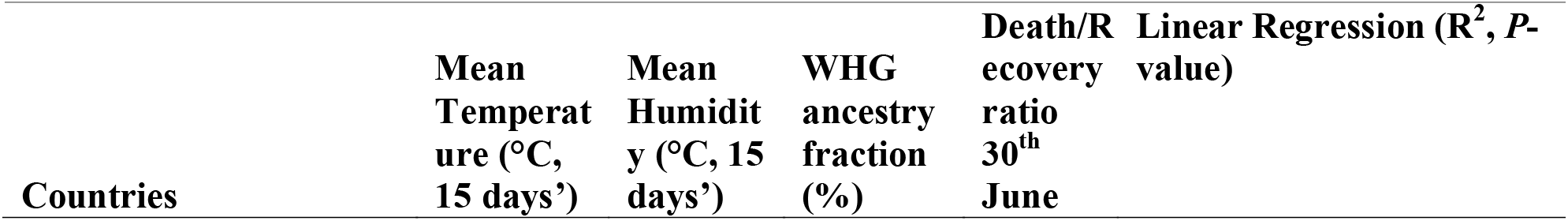
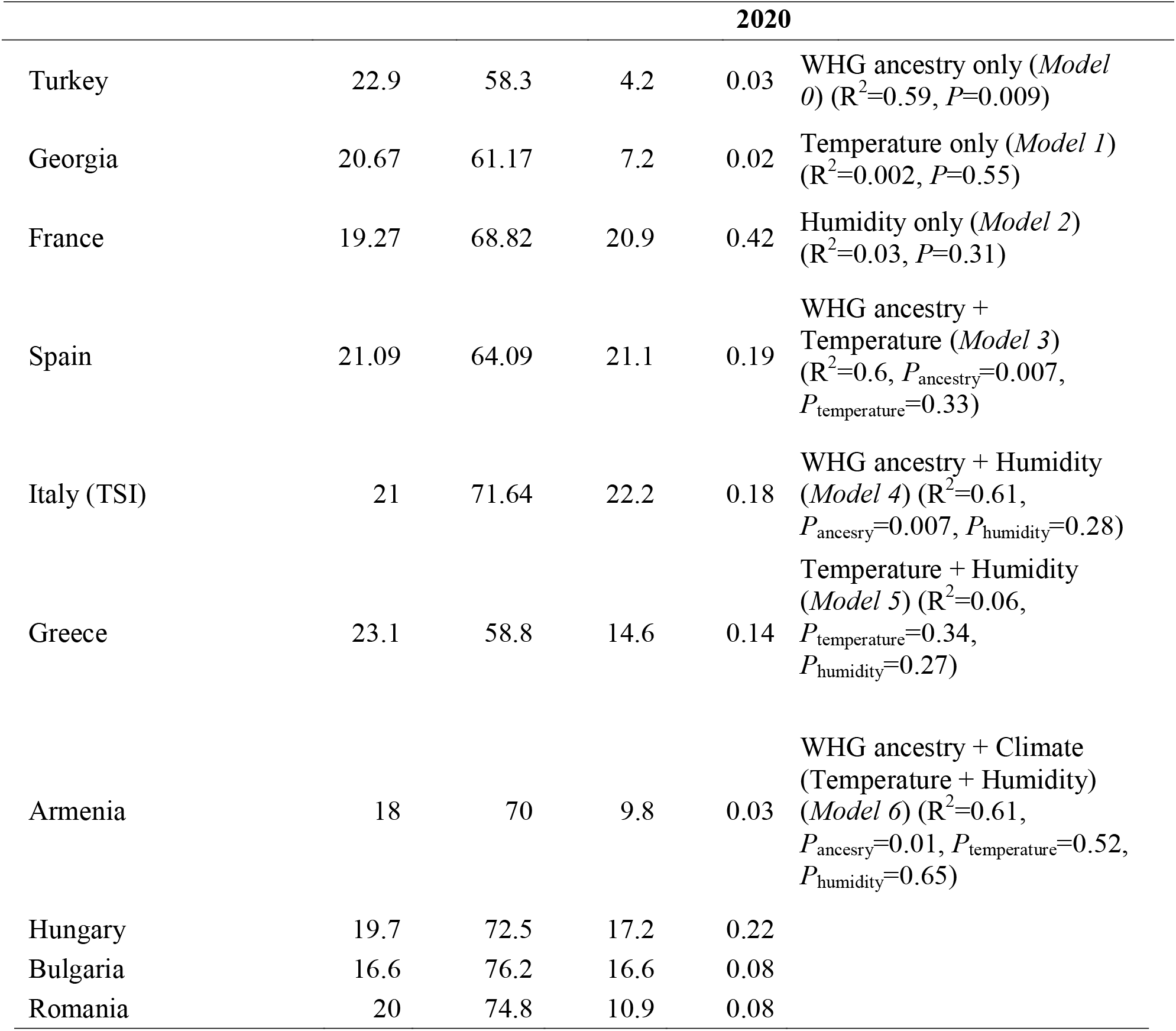
West European hunter gatherers (WHGs) ancestry fractions, mean temperature (15 days), mean humidity (15 days), and COVID-19 death/recovery ratios of 10 European countries employed in the study for reanalysis (data obtained for 30^th^ June 2020)

| Countries | Mean Temperature (°C, 15 days') | Mean Humidity (°C, 15 days') | WHG ancestry fraction (%) | Death/R recovery ratio 30<sup>th</sup> June | Linear Regression ($R^2$, $P$-value) |
| --- | --- | --- | --- | --- | --- |

| 2020 |  |  |  |  |  |
| --- | --- | --- | --- | --- | --- |
| Turkey | 22.9 | 58.3 | 4.2 | 0.03 | WHG ancestry only (*Model 0*) ($R^2=0.59$, $P=0.009$) |
| Georgia | 20.67 | 61.17 | 7.2 | 0.02 | Temperature only (*Model 1*) ($R^2=0.002$, $P=0.55$) |
| France | 19.27 | 68.82 | 20.9 | 0.42 | Humidity only (*Model 2*) ($R^2=0.03$, $P=0.31$) |
| Spain | 21.09 | 64.09 | 21.1 | 0.19 | WHG ancestry + Temperature (*Model 3*) ($R^2=0.6$, $P_{\text{ancestry}}=0.007$, $P_{\text{temperature}}=0.33$) |
| Italy (TSI) | 21 | 71.64 | 22.2 | 0.18 | WHG ancestry + Humidity (*Model 4*) ($R^2=0.61$, $P_{\text{ancestry}}=0.007$, $P_{\text{humidity}}=0.28$) |
| Greece | 23.1 | 58.8 | 14.6 | 0.14 | Temperature + Humidity (*Model 5*) ($R^2=0.06$, $P_{\text{temperature}}=0.34$, $P_{\text{humidity}}=0.27$) |
| Armenia | 18 | 70 | 9.8 | 0.03 | WHG ancestry + Climate (Temperature + Humidity) (*Model 6*) ($R^2=0.61$, $P_{\text{ancestry}}=0.01$, $P_{\text{temperature}}=0.52$, $P_{\text{humidity}}=0.65$) |
| Hungary | 19.7 | 72.5 | 17.2 | 0.22 |  |
| Bulgaria | 16.6 | 76.2 | 16.6 | 0.08 |  |
| Romania | 20 | 74.8 | 10.9 | 0.08 |  |

We used *qpAdm* [20] implemented in AdmixTools v5.1 [21,22] to estimate ancestry proportions in the European genomes originating from a mixture of ‘reference’ populations by utilizing shared genetic drift with a set of ‘outgroup’ populations. 14 ancient genomes namely *Luxembourg_Loschbour.DG, Luxembourg_Loschbour_published.DG, Luxembourg_Loschbour, Iberia_HG* (N=5), *Iberia_HG_lc, Iberia_HG_published, LaBrana1_published.SG, Hungary_EN_HG_Koros* (N=2), *Hungary_EN_HG_Koros_published. SG* were grouped together as West European Hunter-Gatherers (WHGs); three ancient genomes namely *Russia_EHG, Russia_HG_Samara* and *Russia_HG_Karelia* (N=2) were grouped together as Mesolithic hunter-gatherers of Eastern Europe (EHGs); two ancient genomes: *KK1.SG, SATPSG* were grouped as Caucasus hunter-gatherers (CHGs) and *Sweden_HG_Motala* (N=8) were renamed as Scandinavian Hunter-Gatherers (SHGs) in *qpAdm* analysis. We tried to model modern-day Europeans with different combinations of Neolithic Near-East populations such as Neolithic Iranians *(Iran_GanjDareh_N)*, Neolithic Anatolians *(Anatolia_N)* and Natufians *(Israel_Natufian)* alongside various combinations of ancient European Hunter Gatherer populations (WHG, EHG, CHG and SHG) as mentioned in Lazaridis et al. (2016) [23]. We inferred that modern-day Europeans could be best modelled as a combination of three source populations namely EHGs, WHGs and Neolithic Iranians *(Iran_GanjDareh_N)* as *Left* (EHG, WHG, Iran_GanjDareh_N). We used a mixture of eight ancient and modern-day populations comprising of *Ust Ishim, MA1, Kostenki14, Han, Papuan, Chukchi, Karitiana*, Mbutiasour ‘*Right*’ outgroup populations (O8).

Pearson’s correlation coefficient (r) between various ancestry proportions of European genomes and COVID-19 death/recovery ratio was calculated and its significance was statistically evaluated using GraphPad Prism v8.4.0, GraphPad Software, San Diego, California USA [24]. Additionally, we developed several linear regression models with different combinations of WHG ancestry fraction (x_1_) alongside climatic factors such as 15 days’ mean temperatures (x_2_) and 15 days’ mean humidity (x_3_) of the ten European countries employed in this study to statistically evaluate their impact on COVID-19 death/recovery ratio (y) using *glm* function implemented in R v3.6.3 (Table 1, Supplemental Table 1). One-tailed tests were performed considering the alternative hypothesis of positive association of x_1_, x_2_ and x_3_ with y (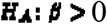). The best model was determined by likelihood ratio test (LRT), employing *lrtest* function, implemented in R package ‘*epicalc’* Temperature and humidity data were obtained from Time and Date.com [25], which garners meteorological data from leading meteorological institutes such as The World Meteorological Organization [26] and Meteorological Assimilation Data Ingest System (MADIS) [27]. Ancestry and COVID-19 death/recovery ratio maps were mapped by country using QGIS version 3.12.3 [28].

Notably, the death/recovery ratio was used in this study as the test statistic as it can potentially best reflect the extent to which individuals have succumbed versus those recovering owing to COVID-19 infection. We believe that this ratio likely best reflects host immune responsiveness against viral assault, especially since it does not take into account the number of active cases and statistical biases arising due to population size, spread of the disease and under-reporting/under-testing of COVID-19 cases.

### Genome-wide association analyses (GWAS)

Genome wide association study (GWAS) was performed to identify novel genetic variants that show significant variation between genomes of European and East Asian ancestries. In GWAS, European genomes (N=753) with higher death/recovery ratio (Cases) were compared against Eastern Asian genomes (N=838) with low death/recovery ratio (Controls). To this end, standard case–control-based association analyses was performed in PLINK v1.9 using --assoc command. A sorted list of association results will be generated using --adjust flag to the --assoc command. Chi square test, implemented in PLINK –assoc command, was performed separately for all 597,573 SNPs to statistically assess their significance and for stringency P <0.01 was considered significant. A Manhattan plot was generated in Haploview [29] by plotting Chi square values of all assessed SNPs to identify the SNPs that are likely associated with COVID-19 manifestations. Highly significant SNPs were annotated using SNPnexus web-based server [30].

## Results

### Ancestry proportions in European genomes and COVID-19 manifestations

We modelled all Europeans as a combination of three source populations namely EHGs, WHGs and Neolithic Iranians (see Methods) in *qpAdm* analysis. Among 10 European populations employed in this study, GBR (British in England and Scotland) genomes were found to have the highest WHG ancestry proportions (26.1%) (Fig. 1a, Table 1). Notably, among the European populations evaluated, COVID-19 death/recovery ratio till 5^th^ April 2020, was the highest among the British people: death/recovery ratio=31.94 (Fig. 1b, Table 1). However, we omitted GBR genomes in our reanalysis due to absence of updated COVID-19 recovery information from UK. In contrast, Russian and Finnish populations which could not be modelled with WHG ancestry so far appear to have less detrimental COVID-19 manifestations with 0.02 and 0.05 death/recovery ratio, respectively (Fig. 1a and 1b). Consistent with this, we found significant positive correlation (Pearson’s Correlation; r=0.58; *P*=0.03) between WHG ancestry fraction and COVID-19 death/recovery ratio. The association discernibly increased (r=0.77, *P*=0.009) upon reanalysing the data, omitting countries with small sample sizes and adding those that form a bridge between Europe and Asia.

**Fig. 1:**
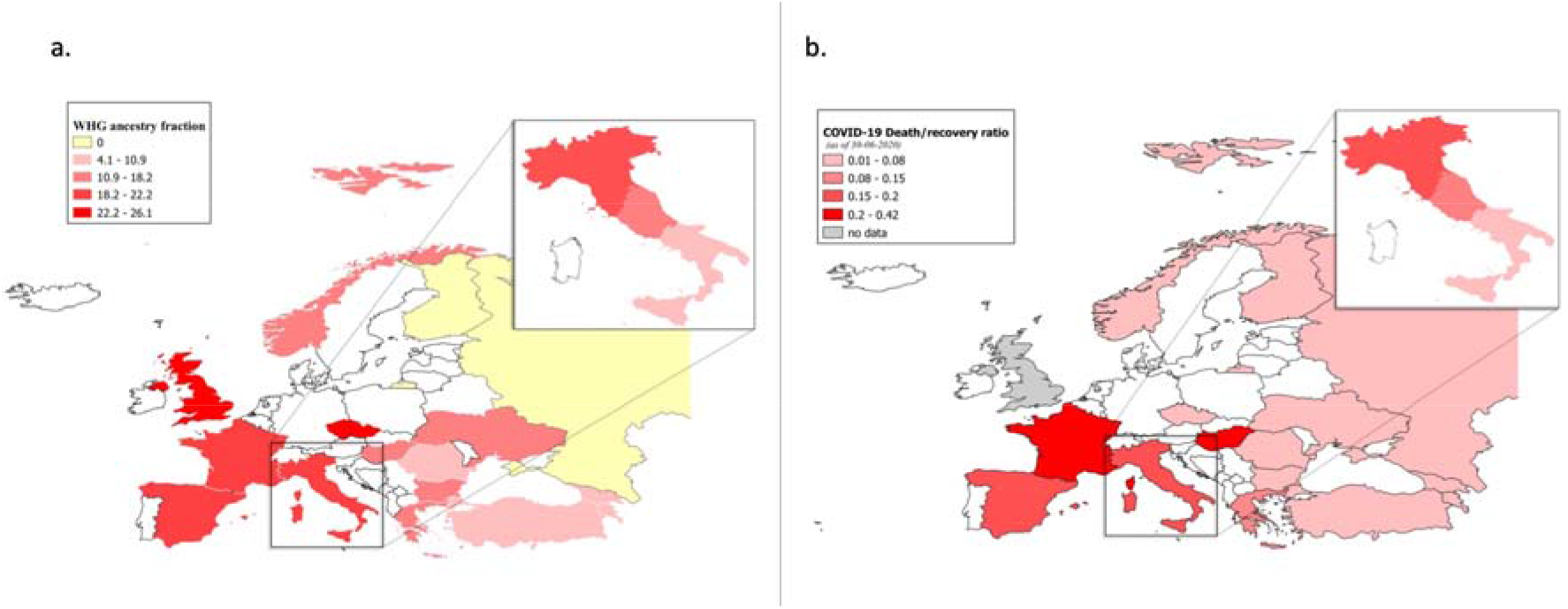
Maps showing WHG ancestry fractions (a) and COVID-19 death/recovery ratio as of 30^th^ June 2020 (b) in various European countries. Maps were plotted in QGIS software (http://qgis.osgeo.org). To show gradient decrease in WHG ancestry fractions, WHG ancestry proportion in Central Italy (a: inset) was calculated by taking an average of WHG ancestry proportions of Northern (23.1%) and Southern (9.2%) Italy. Finland and Russia which could not be modelled to WHG ancestry are shown in yellow (a) and since current COVID-19 recovery rate in UK is unknown, it is shown in grey (b).

Congruent with correlation results, we found significant linear positive association between WHG ancestry fraction and COVID-19 death/recovery ratio (Model 0) (R^2^=0.34, *P* =0.039) (Table 1). However, we did not find any association between 15 days’ mean temperature (Model 1) and humidity (Model 2) with COVID-19 death/recovery ratio (*P* =0.49 and 0.29 respectively) (Table 1). Moreover, we found significant association between the combination of WHG ancestry and temperature (Model 3), WHG ancestry and humidity (Model 4), and WHG ancestry and a combination of both climatic factors (temperature and humidity, Model 6) with COVID-19 death/recovery ratio (R^2^=0.33, 0.39 and 0.4 respectively) (Table 1). However, in all cases the significance of the test was driven by the WHG ancestry fraction alone (*P*=0.04, 0.04 and 0.05 for Model 3, Model 4 and Model 6 respectively) and not by the climatic factors (*P*_temperature_=0.54, *P*_humidity_=0.23, and *P*_temperature_=0.61 and *P*_humidity_=0.23 for Model 3, Model 4 and Model 6 respectively). Consistent with this, Likelihood Ratio Test (LRT) determined Model 0 to be the best model, indicating that the extent of COVID-19 disease severity can be best explained by variability in WHG ancestry fractions among European populations. Model 0 was found to be highly significantly better than Model 1 (Temperature alone) and Model 2 (Humidity alone) *(P* ≈ 0) in recapitulating the variability of COVID-19 death/recovery ratio among the study populations, suggesting that climatic factors such as colder temperature and variation in humidity alone cannot explain the variability of COVID-19 disease in European countries. Further, the combination of ancestry and temperature, ancestry and humidity (Model 3 and 4 respectively), and the combination of ancestry and climate (temperature + humidity) (Model 6) failed to explain variabilities in COVID-19 death/recovery ratio better than ancestry alone (Model 0) (*P* =0.88, 0.36 and 0.61 respectively). This largely eliminates the possibility of regional climate playing major role in the severity of COVID-19 presentation.

The association between WHG ancestry fraction (Model 0) and COVID-19 death/recovery ratio discernibly increased during reanalysis (R^2^=0.59, *P*=0.009) (Table 2). Consistent with the initial analysis, we did not find any association between 15 days’ mean temperature (Model 1) and humidity (Model 2) with COVID-19 death/recovery ratio (*P* =0.55 and 0.31 respectively) and found significant association between the combination of WHG ancestry and temperature (Model 3), WHG ancestry and humidity (Model 4), and WHG ancestry and the combination of both climatic factors (temperature and humidity, Model 6) with COVID-19 death/recovery ratio (R^2^=0.6, 0.61 and 0.61 respectively) (Table 2). However, similar to our initial analysis, the significance of the test was driven by the WHG ancestry fraction alone (*P*=0.01, 0.01 and 0.02 for Model 3, Model 4 and Model 6 respectively) and not by the climatic factors (*P*_temperature_=0.33, *P*_humidity_=0.28, and *P*_temperature_=0.48 and *P*_humidity_=0.35 for Model 3, Model 4 and Model 6 respectively) (Table 2). Finally, congruent with our initial analysis, LRT determined Model 0 to be the best model, indicating COVID-19 manifestations can be best explained by variability in WHG ancestry fractions among European populations.

### Genome-wide association analyses (GWAS)

For GWAS, 753 genomes from across Europe with high COVID-19 death/recovery ratio (case) were compared against 838 Eastern Asian genomes with relatively lower COVID-19 death/recovery ratio (control). Out of 597,573 SNPs employed in this study, 385,450 (64.5%) markers revealed highly significant variation (Chi square≥ 6.63; P<0.01) between Europeans and East Asians, indicating the discernible differences in the genetic tapestry of the two populations (Fig. 2). For stringency, we annotated only the top 10,000 ranked SNPs (N=10,011, Chi square≥ 739; P<9.69×10^-163^) using SNPnexus web-based server [30] (Supplemental Table 2). Among top 10,000 ranked SNPs, 404 were associated with host immune responsiveness including interferon (IFN) stimulated antiviral response, Interferon-stimulated gene 15 *(ISG15)* mediated antiviral mechanism and 2′-5′ oligoadenylate synthase (OAS) mediated antiviral response. These 10,000 SNPs are linked to 161 genes associated with immune system regulation including those related to innate immunity, antiviral response and receptors (eg. *EDAR)* (Supplemental Table 3). Our results indicate that pathways such as those involved in immunoregulatory interactions between a lymphoid and a non-lymphoid cell, adaptive and innate immune system, IL-6 dependent immune responses, and complement cascades show significant variation between genomes of East Asian and European ancestry. Among these, type-I interferon and IL-6 dependent inflammatory immune responses have been shown to be associated with SARS-CoV-2 infection [31].

**Fig. 2:**
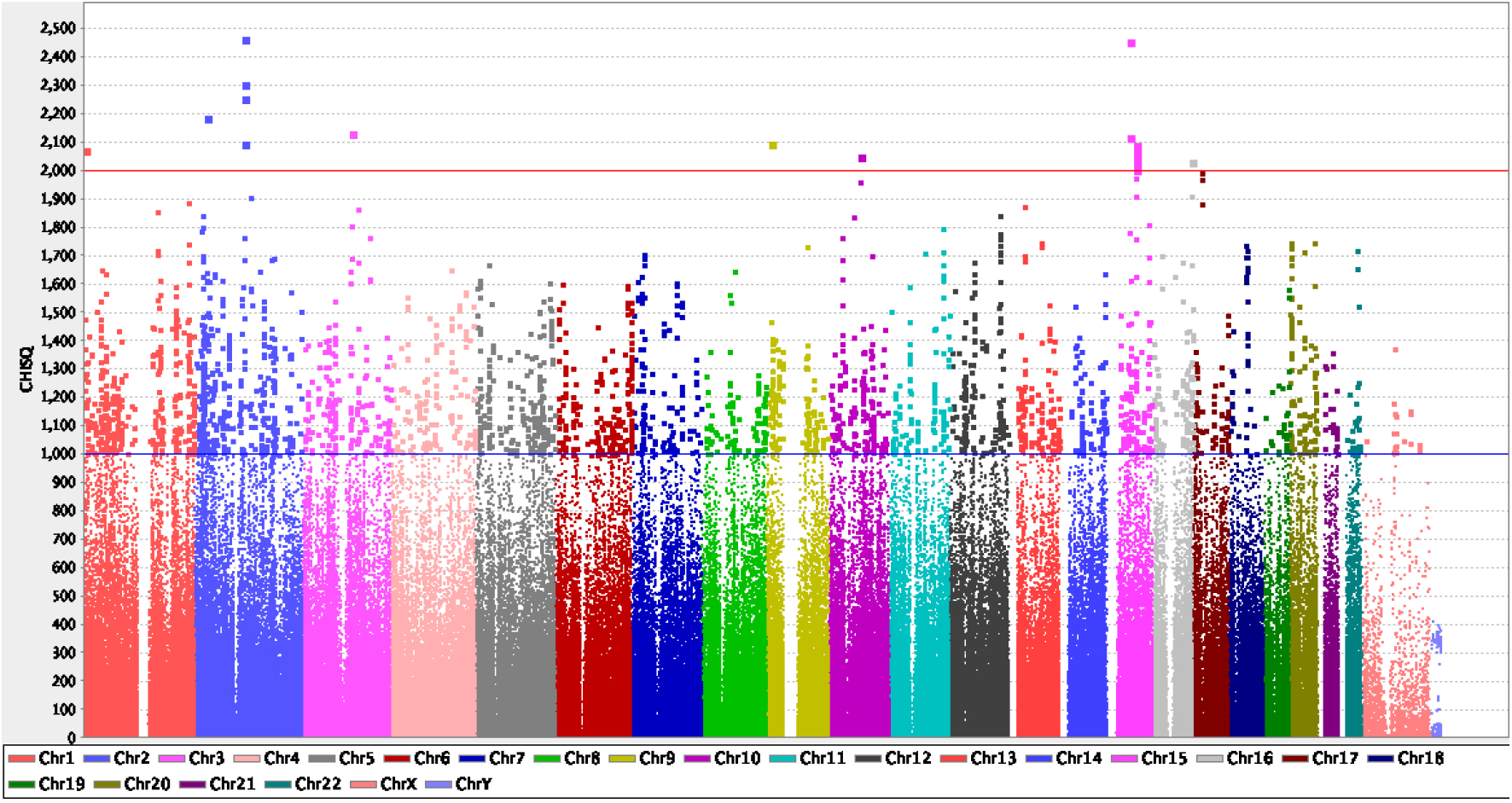
Manhattan Plot. X-axis represents chromosomes (chr 1 to chr Y). SNPs present in the chromosomes are designated with dots. The chi square values are plotted in Y-axis. GWAS with 753 European genomes were compared to 838 Eastern Asian genomes. Out of 597,573 SNPs employed, 385,450 (64.5%) markers revealed highly significant variation between Europeans and East Asians. The SNPs with chi square values >1000 are indicated with the blue line and those with chi square value >2000 are indicated with the red line. While 18 SNPs showed chi square values >2000, 2,992 SNPs had chi square values >1000.

Furthermore, the SNPs with the highest Chi square values (≥1500; *P*≈0) were found to be associated with immune system related pathways such as Senescence-Associated Secretory Phenotype (SASP), Interleukin-4 and Interleukin-13 signaling, TNFR2 non-canonical NF-kB pathway and Class I MHC mediated antigen processing and presentation (Supplemental Table 4). Among these, Interleukin mediated ‘cytokine storm’ has been shown to be associated with pulmonary inflammation among COVID-19 patients [32], and the genetic variation in Class I MHC has been associated with severity of COVID-19 manifestations [9].

Finally, six immune system related SNPs (rs2243250, rs1800872, rs1800896, rs1544410, rs1800629 and rs1805015) that display significant variability between Europeans and East Asians (Odds Ratio>2, Chi square>40, *P*<10^-10^) are likely associated with the development of immune responses in the first year of life contingent upon the gene-environment interactions [33].

## Discussion

The novel coronavirus SARS-CoV-2 infection has shown discernible worldwide variability. While in some areas affected individuals are recovering relatively quicker, in others, recovery times have been comparatively longer, and mortality high. Here, we sought to investigate the likely association between genetic ancestry determinants and the extent of COVID-19 manifestations employing Europeans as a case study.

We found a significant positive correlation (r=0.58, P=0.03) between West European hunter gatherers (WHG) ancestral fractions and COVID-19 death/recovery ratio which discernibly increased (r=0.77, P=0.009) upon reanalyses as of 30^th^ June 2020, removing countries with small sample sizes and including those from intermediate geographical locations between Europe and Asia, such as Georgia, Armenia and Turkey. We identified 404 immune response related SNPs by comparing publicly available data from 753 genomes corresponding to various European countries against 838 genomes from various Eastern Asian countries using GWAS. We found that SNPs associated with immune-system related pathways such as interferon stimulated antiviral response, adaptive and innate immune system and IL-6 dependent immune responses show significant differences in allele frequencies [Chi square values (≥1500; *P*≈0)] between Europeans and East Asians.

### The European story

Europe has been severely impacted owing to the ongoing COVID-19 pandemic. Our findings revealed a significant positive correlation between WHG-related ancestry and COVID-19 death/recovery ratio (*P_initial analysis_*=0.03, *P_reanaiysis_*=0.009). Further, we found significant linear positive association between WHG ancestry fraction and COVID-19 death/recovery ratio (R^2^*_initial analysis_* = 0.34, *P_initial analysis_* = 0.039, R^2^*_reanalysis_* = 0.59, *P_reanalysis_* = 0.009). Additionally, we found that the regional climate alone is unlikely to play a major role in determining the severity of COVID-19 disease. Previously it has been shown that European genomes evolved uniquely with regards to immune responsiveness, particularly pertaining to responses against viral infections [8]. Admixture with Neanderthals is believed to have introduced unique regulatory variants into European genomes, which potentially regulated immune responsiveness in European populations [7].We surmise that the European genome architecture has been extensively modulated by the complex origin and migration history of modern-day Europeans during Paleolithic, Mesolithic and Neolithic periods that led to the introduction of novel variants in European genomes.

Modern-day European genomic diversity has been thought to be shaped by variable proportions of local hunter-gatherer ancestry (WHGs) [34]. While Neolithic, Iberian genomes (modern day Spanish and Portuguese populations) revealed widespread evidence of WHG admixture (10-27%), Neolithic German populations (Linearb and keramik– LBK culture) revealed ~4-5% of the same [34]. Notably the variation in WHG ancestry fractions among German and Spanish people correlated with the observed variabilities in COVID-19 death/recovery ratio in these two countries; German populations with discernably lower WHG fractions have a current death/recovery ratio of 0.05, however, in Spain the ratio is 0.19, which corresponded to high WHG fraction among Spanish populations. A similar correlation between WHG ancestry and the extent of COVID-19 manifestations was observed for the UK, where GBR genomes depicted high WHG ancestry fractions (>25%) and was correlated with high COVID-19 death/recovery ratio in the country (31.94). We note here that due to unavailability of official COVID-19 recovery information from the UK since April 2020, we were unable to include UK in our reanalyses.

Italy, one of the worst hit European countries during the COVID-19 pandemic showed significant nationwide variation in the severity of disease manifestations (death/recovery ratio=0.18) with a clear gradient in COVID-19 mortality rates from north to south. While COVID-19 death/recovery ratio in Northern Italy is 0.19, the ratio is 0.15 in Central Italy and 0.11 in Southern Italy. Interestingly, these numbers are consistent with WHG fractions among Italian genomes. While Northern Italian genomes have ~23.1% WHG ancestry fractions, the fraction reduces to 9.2% among Southern Italians (Fig. 1a and 1b).

In contrast, although COVID-19 cases have been rising rapidly in Russia, their death/recovery ratio (0.02) has remained significantly lower than most European countries. We assessed whether this is owing to their genomic make-up; all modern-day Russians originated from two groups of East Slavic tribes: Northern and Southern [35] and have 0% WHG ancestry fraction. They are genetically similar to modern-day other Slavic populations such as Belarusians [35,36]. Consistent with this we observed significantly low COVID-19 death/recovery ratios in both Russia (0.02) and Belarus (0.01). In addition, COVID-19 manifestations were found to be far less detrimental in the Central Asian countries part of the erstwhile Soviet Union (USSR) such as Kazakhstan (death/recovery ratio=0.01), Kyrgyzstan (0.02), Tajikistan (0.01), and Uzbekistan (0.005).

Finally, Finnish genomes that were shaped predominantly by migrations from Siberia ~3500 years ago [36] with very small WHG ancestral fractions, also have a discernibly smaller COVID-19 death/recovery ratio (0.05) compared to neighboring Sweden, where the ratio (0.88) is among the highest in world as of 31^st^ May, 2020, after which no official recovery information was available [2]. We note here that hunter gatherer genomes from Sweden have shown to form a WHG cluster including Loschbour and La Braña2, indicating high affinity of Swedish genomes towards WHG ancestry [37].

Historically, Turkey has been a crossroad of major population migrations across Eurasia and served as a socio-cultural bridge between Europe and Asia. A whole genome-based study revealed that while the modern-day Turkish population are genetically similar to Southern European populations such as Italians, most Turkish genomes show signatures of recent genetic contribution from ancestral East Asian populations [38]. On the other hand, modern-day Armenians derived 29% of their European ancestry from Tyrolean Iceman [39], a 5300-year-old Copper age individual, discovered along Tisenjoch Pass of the Ötztal Alps [40]. Consequently, WHGs who contributed discernible ancestry fractions to most Europeans, provided very low fraction (<10%) of the same in the ancestries of Near Easterners (Turkey, Georgia and Armenia) [37]. Consistent with this, COVID-19 death/recovery ratio in Turkey, Georgia and Armenia remained discernibly lower compared to most European countries (0.03, 0.02 and 0.03 respectively), further underscoring the importance of WHG-related ancestral components in determining the severity of COVID-19 manifestations.

Overall, our results have revealed a clear association between WHG ancestry fractions and the acuteness of COVID-19 manifestations suggesting the presence of unique underlying genetic variants in European genomes that maybe correlated with increased disease susceptibility and immune responsiveness. It is noteworthy that despite being better equipped in terms of medical facilities and resources for testing and providing care to COVID-19 affected individuals, most western and central European countries have struggled substantially with significantly higher death/recovery ratios, as compared to many Asian countries that are experiencing an increasing burden of patients with chronic underlying conditions necessitating efficient health care systems with integrated care [41]. This further illuminate that the intrinsic host genetic make-up as determined by ancestry is a major determinant of the severity of COVID-19 disease.

### Intrinsic ‘protection’ for East Asians?

As noted above, significant variation in allele frequencies of *ACE2* encoding for the host cell receptor of SARS-CoV-2 has been reported between people of East Asian and European ancestries [6]. The authors of this study speculated that the differences in allele frequencies of *ACE2* coding variants were likely associated with variable expression of ACE2 in tissues. Additionally, they reported that most eQTL variants identified by them had discernibly higher allele frequencies among East Asian populations compared to Europeans, which may result in differential susceptibility towards SARS-CoV-2 infection in these populations under similar conditions. Consistent with these findings we identified 404 SNPs that are associated with host immune response, such as Interferon (IFN) stimulated antiviral response, Interferon-stimulated gene 15 (ISG15) mediated antiviral mechanism and 2′-5′ oligoadenylate synthase (OAS) mediated antiviral response that showed large differences in allele frequencies between Europeans and East Asians. The genetic differences between East and Southeast Asians with Europeans can be largely attributed to their distinctive ancestral origins. Most East Asians derive their ancestry from Mongolian hunter-gatherers who dispersed over Northeast Asia 6000-8000 years ago [42]. Similarly, Southeast Asians exhibit a mixture of East Asian ancestry (Southern Chinese agriculturalist) and a diverged form of Eastern Eurasian hunter-gatherer ancestry (EHGs) [43]. People with similar ancestry can be found as far south as Indonesia [43]. As noted above EHG ancestry fraction does not appear to be significantly associated with COVID-19 manifestations. In congruence with their unique ancestral make-up, all East and Southeast Asian countries exhibit discernibly low COVID-19 death/recovery ratio compared to most west and central European countries (Japan=0.06, South Korea=0.02, Hong Kong=0.006, Taiwan=0.01, Thailand=0.02, Malaysia=0.01 and Singapore=0.0007). Notably, in mainland China, where the novel corona virus originated, COVID-19 death/recovery ratio has remained discernibly low (0.06). Overall, our results indicate that people of East and Southeast Asian ancestry appear to be intrinsically protected against the most debilitating effects of SARS-CoV-2 infection.

### India: a variegated canvas for COVID-19

Indians have a long and complex history of admixture between immigrant gene-pools originating primarily in West Eurasia, Southeast Asia and the Indian hunter-gatherer lineage with close genetic proximity to the present-day Andamanese people (Ancient Ancestral South Indians: AASI) who likely arrived in India through the “southern exit” wave out of Africa [44]. People of AASI ancestry admixed with an undivided ancient Iranian lineage that subsequently split and lead to the formation of early Iranian farmers, herders, and hunter gatherers approximately in the 3^rd^ millennium BCE. This gene pool is referred to as the ‘Indus Periphery’ gene pool [44,45], which is thought to be the major source of subsequent peopling of India. Modern-day Indian genome is composed of largely four ancestral components: Ancestral North Indian (ANI), Ancestral South Indian (ASI), Ancestral Tibeto-Burman (ATB) and Ancestral Austro-Asiatic (AAA) [46]. ANI and ASI gene pools likely arose around 2^nd^ millennium BCE during the decline of Indus Valley Civilization (IVC), which prompted multiple waves of migrations across the India. The southward migration of Middle to Late Bronze Age people from Steppe (Steppe MLBA) into India is thought to have coincided with the decline of IVC. It is speculated that the people of Indus-Periphery-related ancestry, while migrating northward, admixed with Steppe MLBA immigrants to form the ANI, while the others, who migrated southward and eastward admixed with AASI and formed the ASI [45]. Austroasiatic speakers originated in Southeast Asia and subsequently migrated to India during the Neolithic period. The admixture between local Indians and incoming Southeast Asians took place ~2000-3800 years ago giving rise to the AAA ancestry. Notably, the Indian population(s) with whom the incoming Southeast Asians mingled were AASI related with little to no West Eurasian ancestry fraction [47]. Finally, it has been showed that the Tibeto-Burmans (ATBs) derived their ancestry through admixture with low-altitude East Asians who migrated from China and likely across Northern India or Myanmar [48] leading to the high genomic proximity between Indians of ATB ancestry and East Asians.

We believe that the severity of COVID-19 disease presentation is likely to vary appreciably across the Indian populations. The genomic proximity between East Asians and Indians mostly from Northeast India with prominent ATB ancestry, is reflected through lower COVID-19 mortality rates from this region with Manipur, Mizoram, Nagaland and Sikkim not registering any COVID-19 related death as of 30^th^ June 2020 [19]. Notably the death/recovery ratio in the largely ATB dominated region of Ladakh, India is 0.001, indicating almost complete recovery so far amidst COVID-19 infected individuals from this region.

Indian states with large number of indigenous tribal population with high prevalence of AAA ancestry and/or with large fractions of AASI-related ancestry, eg. Andaman and Nicobar Islands, Chhattisgarh, Jharkhand and Orissa have registered very few COVID related deaths till date (death/recovery ratio 0, 0.005, 0.008, 0.004 respectively). Furthermore, Tamil Nadu, despite ranking among the top three worst affected states in India, in terms of the numbers of COVID-19 positive cases have registered significantly lower COVID-19 death/recovery ratio (0.02) that could be attributed to discernible fractions of AASI-related ancestry among the people from this area. This indicates that Indians with predominant AAA and AASI related ancestry may likely be less severely affected against the most detrimental effects of COVID-19 infection. On the other hand, states with predominant ANI ancestry such as Gujarat, Maharashtra, Madhya Pradesh and Delhi have shown discernibly higher COVID-19 mortality rates (0.08, 0.08, 0.06 and 0.05, respectively).

We also note that approximately 83% of WHG ancestry has been associated with mitochondrial DNA U haplogroup [49,50]. Overall, 13.1% Indians belong to U haplogroup and are thus associated with WHG related ancestry [51]. And the U haplogroup frequency shows discernible variation among Indian populations, while 23.3% North Indians belong to U haplogroup, 10.3% South Indians belong to the same [51]. U haplogroup also shows discernible variation across Indian societal structure, while 15% of Indian upper castes belong to U haplogroup, 8% of Indian tribal populations (mostly of AAA ancestry) have the same [52]. Taken together, overall ~8-19% Indians are likely to have WHG related ancestry. Notably, our previous whole genome-based study found most Gujarati people to have ~3-4% WHG related ancestry [53], similar to what is observed for Turkish Jews in the current study. Our interrogation of worldwide populations suggests that the WHG ancestral fraction is likely associated with acute COVID-19 manifestations and is predictive of debilitating effects of COVID-19 infection among Indian populations with substantial fractions of WHG ancestry. Overall COVID-19 manifestations in India is likely to be somewhat intermediate to that observed in Europe and East Asia. This is also supported by the recent finding that allele frequencies of *ACE2* variants among Indians are intermediate between East Asians and Europeans [6].

### Limitations and conclusion

The present study shines light of underlying genetic signatures that may be associated with disparate COVID-19 severity and manifestations in worldwide populations. Nevertheless we note that the current work has been performed using publicly available genomic data and a more robust understanding in this regard will emanate from sequencing/genotyping endeavours for COVID-19 patients across the spectrum of varied nationalities/ancestries and geographical locations, including individuals with mild to moderate symptoms, severe manifestations and death. We further note that since the current analyses is performed using pre-existing genomic data, there is a disparity in number of individuals sequenced among various populations which might influence the analyses in terms of statistical power and errors.

We also note that populations from most European countries have higher mean ages compared to India, which may accentuate mortality rates among Europeans. However, as reported recently age alone may not suffice in exacerbating death/recovery ratios, and underlying health conditions such as cardiovascular disease, chronic kidney disease, hypertension, chronic respiratory disease, diabetes, cancers, HIV/AIDS, tuberculosis etc., many of which are already described as crucial comorbidity factors for COVID-19 may modify disease manifestations and prognosis in patients [54]. It is imperative to consider here that although the mean population age is lower in India compared to most European countries, 7.7% Indians suffer from diabetes [55], 25.3% suffer from hypertension [56] and approximately 14.5% suffer from some form of respiratory disorders linked to air pollution [57], all of which are underlying health conditions that are reported to cause higher COVID-19 death/recovery ratio [46]. Therefore, lower death/recovery ratio among Indians, particularly in specific populations compared to most European countries highlights the role of host genetic constitution in influencing COVID-19 disease progression.

Finally, several factors such as demographics (eg. population size and density), healthcare facilities and hospital infrastructure, administrative preparedness and implementation of social isolation and lockdowns were beyond the scope of the current study but may influence the death/recovery metric of a country. Taken together our results strongly advocate for the adoption of a rigorous worldwide population genetics driven approach to expand and substantiate our current knowledge of SARS-CoV-2 infection, as well as facilitate the development of population specific therapeutics to mitigate this worldwide challenge.

## Data Availability

The current study employed publicly available data, which is freely available through the database of Dr. David Reich, Harvard Medical School (https://reich.hms.harvard.edu/datasets).

## References

1. Laboratory testing of 2019 novel coronavirus (2019-nCoV) in suspected human cases: interim guidance, 17 January 2020 [Internet]. [cited 2020 Apr 9]. Available from: https://appsx.who.int/iris/handle/10665/330676

2. Dong E, Du H, Gardner L. An interactive web-based dashboard to track COVID-19 in real time. Lancet Infect Dis 2020 20(5):533–534 PMID:32087114

3. Verity R, Okell LC, Dorigatti I, Winskill P, Whittaker C, Imai N, et al. Estimates of the severity of coronavirus disease 2019: a model-based analysis. Lancet Infect Dis 2020 20(6):669–677. PMID:32240634

4. Zhou P, Yang X-L, Wang X-G, Hu B, Zhang L, Zhang W, et al. A pneumonia outbreak associated with a new coronavirus of probable bat origin. Nature 2020 Mar;579(7798):270–273. PMID:32015507

5. Lu R, Zhao X, Li J, Niu P, Yang B, Wu H, et al. Genomic characterisation and epidemiology of 2019 novel coronavirus: implications for virus origins and receptor binding. The Lancet 2020 Feb;395(10224):565–574. PMID:32007145

6. Cao Y, Li L, Feng Z, Wan S, Huang P, Sun X, et al. Comparative genetic analysis of the novel coronavirus (2019-nCoV/SARS-CoV-2) receptor ACE2 in different populations. Cell Discov 2020 Dec;6(1):11. PMID:32133153

7. Stawiski EW, Diwanji D, Suryamohan K, Gupta R, Fellouse FA, Sathirapongsasuti F, Liu J, Jiang YP, Ratan A, Mis M, Santhosh D. Human ACE2 receptor polymorphisms predict SARS-CoV-2 susceptibility. bioRxiv. 2020 Jan 1.

8. Li G, Fan Y, Lai Y, Han T, Li Z, Zhou P, et al. Coronavirus infections and immune responses. J Med Virol 2020 Apr;92(4):424–432. PMID:31981224

9. Nguyen A, David JK, Maden SK, Wood MA, Weeder BR, Nellore A, Thompson RF. Human leukocyte antigen susceptibility map for SARS-CoV-2. Journal of virology. 2020 Jun 16;94(13):e00510–20. PMID:32303592.

10. Kuo CL, Pilling LC, Atkins JL, Masoli JA, Delgado J, Kuchel GA, et al. APOE e4 genotype predicts severe COVID-19 in the UK Biobank community cohort. J Gerontol A Biol Sci Med Sci 2020 May 26;glaa131. PMID:32451547

11. Quach H, Rotival M, Pothlichet J, Loh Y-HE, Dannemann M, Zidane N, et al. Genetic Adaptation and Neandertal Admixture Shaped the Immune System of Human Populations. Cell 2016 Oct 20;167(3):643–656.e17. PMID:27768888

12. Mathieson I, Alpaslan-Roodenberg S, Posth C, Szécsényi-Nagy A, Rohland N, Mallick S, et al. The genomic history of southeastern Europe. Nature 2018 Mar;555(7695):197–203. PMID:29466330

13. Olalde I, Brace S, Allentoft ME, Armit I, Kristiansen K, Booth T, et al. The Beaker phenomenon and the genomic transformation of northwest Europe. Nature 2018 Mar;555(7695):190–196. PMID:29466337

14. Olalde I, Mallick S, Patterson N, Rohland N, Villalba-Mouco V, Silva M, et al. The genomic history of the Iberian Peninsula over the past 8000 years. Science 2019 Mar 15;363(6432):1230–1234. PMID:30872528

15. Kachuri L, Francis SS, Morrison M, Bossé Y, Cavazos TB, Rashkin SR, et al. The landscape of host genetic factors involved in infection to common viruses and SARS-CoV-2. medRxiv. 2020 Jan 1. PMID:32511533

16. David Reich Lab datasets URL https://reich.hms.harvard.edu/datasets [accessed 2020-03-251

17. Price AL, Patterson NJ, Plenge RM, Weinblatt ME, Shadick NA, Reich D. Principal components analysis corrects for stratification in genome-wide association studies. Nat Genet 2006 Aug;38(8):904–909. PMID:16862161

18. Purcell S, Neale B, Todd-Brown K, Thomas L, Ferreira MAR, Bender D, et al. PLINK: A Tool Set for Whole-Genome Association and Population-Based Linkage Analyses. Am J Hum Genet 2007 Sep;81(3):559–575. PMID:17701901

19. India COVID-19 statewise status URL https://www.mygov.in/corona-data/covid19-statewise-status

20. Haak W, Lazaridis I, Patterson N, Rohland N, Mallick S, Llamas B, et al. Massive migration from the steppe was a source for Indo-European languages in Europe. Nature 2015 Jun;522(7555):207–211. PMID:25731166

21. Patterson N, Moorjani P, Luo Y, Mallick S, Rohland N, Zhan Y, et al. Ancient Admixture in Human History. Genetics 2012 Nov;192(3):1065–1093. PMID:22960212

22. Alexander DH, Novembre J, Lange K. Fast model-based estimation of ancestry in unrelated individuals. Genome Res 2009 Sep 1;19(9):1655–1664. PMID:19648217

23. Lazaridis I, Nadel D, Rollefson G, Merrett DC, Rohland N, Mallick S, et al. Genomic insights into the origin of farming in the ancient Near East. Nature. 2016 Aug;536(7617):419–24.

24. Graphpad Prism 8 URL www.graphpad.com

25. Time and Date website URL https://www.timeanddate.com/

26. World Meteorological Organization URL https://public.wmo.int/en

27. Meteorological Assimilation Data Ingest System (MADIS) URL https://madis.ncep.noaa.gov/

28. Team QD. 2016. QGIS geographic information system. Open Source Geospatial Foundation Project. URL http://qgis.osgeo.org

29. Barrett JC, Fry B, Maller J, Daly MJ. Haploview: analysis and visualization of LD and haplotype maps. Bioinformatics 2005 Jan 15;21(2):263–265. PMID:15297300

30. Dayem Ullah AZ, Oscanoa J, Wang J, Nagano A, Lemoine NR, Chelala C. SNPnexus: assessing the functional relevance of genetic variation to facilitate the promise of precision medicine. Nucleic Acids Res 2018 Jul 2;46(W1):W109–W113. PMID:29757393

31. Ramlall V, Thangaraj PM, Tatonetti NP, Shapira SD. Identification of Immune complement function as a determinant of adverse SARS-CoV-2 infection outcome. medRxiv. 2020 Jan 1. PMID:32511494

32. Coperchini F, Chiovato L, Croce L, Magri F, Rotondi M. The cytokine storm in COVID-19: an overview of the involvement of the chemokine/chemokine-receptor system. Cytokine & Growth Factor Reviews. 2020 May 11. PMID:32446778

33. SNPedia URL https://www.snpedia.com

34. Lipson M, Szécsényi-Nagy A, Mallick S, Pósa A, Stégmár B, Keerl V, et al. Parallel palaeogenomic transects reveal complex genetic history of early European farmers. Nature 2017 Nov;551(7680):368–372. PMID:29144465

35. Morozova I, Evsyukov A, Kon–kov A, Grosheva A, Zhukova O, Rychkov S. Russian ethnic history inferred from mitochondrial DNA diversity. Am J Phys Anthropol 2012 Mar;147(3):341–51. PMID:22183855

36. Lamnidis TC, Majander K, Jeong C, Salmela E, Wessman A, Moiseyev V, et al. Ancient Fennoscandian genomes reveal origin and spread of Siberian ancestry in Europe. Nat commun. 2018 Nov 27;9(1):5018. PMID:30479341

37. Lazaridis I, Patterson N, Mittnik A, Renaud G, Mallick S, Kirsanow K, Sudmant PH, Schraiber JG, Castellano S, Lipson M, Berger B. Ancient human genomes suggest three ancestral populations for present-day Europeans. Nature. 2014 Sep;513(7518):409–13. PMID:25230663

38. Alkan C, Kavak P, Somel M, Gokcumen O, Ugurlu S, Saygi C, et al. Whole genome sequencing of Turkish genomes reveals functional private alleles and impact of genetic interactions with Europe, Asia and Africa. BMC genomics. 2014 Dec 1;15(1):963. PMID:25379065

39. Haber M, Mezzavilla M, Xue Y, Comas D, Gasparini P, Zalloua P, et al. Genetic evidence for an origin of the Armenians from Bronze Age mixing of multiple populations. European Journal of Human Genetics. 2016 Jun;24(6):931–6. PMID:26486470

40. Keller A, Graefen A, Ball M, Matzas M, Boisguerin V, Maixner F, Leidinger P, et al. New insights into the Tyrolean Iceman’s origin and phenotype as inferred by whole-genome sequencing. Nat commun. 2012 Feb 28;3(1):1–9. PMID:22426219

41. Tham TY, Tran TL, Prueksaritanond S, Isidro JS, Setia S, Welluppillai V. Integrated health care systems in Asia: an urgent necessity. Clinical interventions in aging. 2018;13:2527. PMID:30587945

42. Wang C-C, Yeh H-Y, Popov AN, Zhang H-Q, Matsumura H, Sirak K, et al. The Genomic Formation of Human Populations in East Asia [Internet]. Genomics; 2020 Mar. [doi: 10.1101/2020.03.25.004606]

43. Lipson M, Cheronet O, Mallick S, Rohland N, Oxenham M, Pietrusewsky M, et al. Ancient genomes document multiple waves of migration in Southeast Asian prehistory. Science 2018 Jul 6;361(6397):92–95. PMID:29773666

44. Shinde V, Narasimhan VM, Rohland N, Mallick S, Mah M, Lipson M, et al. An Ancient Harappan Genome Lacks Ancestry from Steppe Pastoralists or Iranian Farmers. Cell 2019 Oct;179(3):729–735.e10. PMID:31495572

45. Narasimhan VM, Patterson N, Moorjani P, Rohland N, Bernardos R, Mallick S, et al. The formation of human populations in South and Central Asia. Science 2019 Sep 6;365(6457):eaat7487. PMID:31488661

46. Basu A, Sarkar-Roy N, Majumder PP. Genomic reconstruction of the history of extant populations of India reveals five distinct ancestral components and a complex structure. Proc Natl Acad Sci 2016 Feb 9;113(6):1594–1599. PMID:26811443

47. Tätte K, Pagani L, Pathak AK, Koks S, Ho Duy B, Ho XD, et al. The genetic legacy of continental scale admixture in Indian Austroasiatic speakers. Sci Rep 2019 Apr;9(1):6104. PMID:30967570

48. Gnecchi-Ruscone GA, Jeong C, de Fanti S, Sarno S, Trancucci M, Gentilini D, et al. The genomic landscape of Nepalese Tibeto-Burmans reveals new insights into the recent peopling of Southern Himalayas. Sci Rep 2017 Dec;7(1):15512. PMID:29138459

49. Molto JE, Loreille O, Mallott EK, Malhi RS, Fast S, Daniels-Higginbotham J, et al. Complete mitochondrial genome sequencing of a burial from a romano-christian cemetery in the Dakhleh Oasis, Egypt: preliminary indications. Genes. 2017 Oct;8(10):262. PMID:28984839

50. Fu Q, Rudan P, Pääbo S, Krause J. Complete mitochondrial genomes reveal Neolithic expansion into Europe. PloS one. 2012 Mar 13;7(3):e32473. PMID:22427842

51. Kivisild T, Bamshad MJ, Kaldma K, Metspalu M, Metspalu E, Reidla M, et al. Deep common ancestry of Indian and western-Eurasian mitochondrial DNA lineages. Current Biology. 1999 Nov 18;9(22):1331–4. PMID:10574762

52. Karmin M. Human mitochondrial DNA haplogroup R in India: Dissecting the phylogenetic tree of South Asian-specific lineages [Master’s thesis]. University of Tartu; 2005.

53. Das R, Upadhyai P. Investigating the west Eurasian ancestry of Pakistani Hazaras. Journal of genetics. 2019 Jun 1;98(2):43. PMID:31204712

54. Clark A, Jit M, Warren-Gash C, Guthrie B, Wang HH, Mercer SW, et al. Global, regional, and national estimates of the population at increased risk of severe COVID-19 due to underlying health conditions in 2020: a modelling study. Lancet Glob Health. 2020 Jun 15. PMID:32553130

55. Tandon N, Anjana RM, Mohan V, Kaur T, Afshin A, Ong K, et al. The increasing burden of diabetes and variations among the states of India: the Global Burden of Disease Study 1990-2016. The Lancet Global Health. 2018 Dec 1;6(12):e1352–62. PMID:30219315

56. Jose AP, Prabhakaran D. World Hypertension Day: Contemporary issues faced in India. The Indian journal of medical research. 2019 May;149(5):567–570. PMID:31417023

57. Singh V, Sharma BB. Respiratory disease burden in India: Indian chest society SWORD survey. Lung India: Official Organ of Indian Chest Society. 2018 Nov;35(6):459–460. PMID:30381552

